# Causal role of EPA on ischemic heart disease, triglyceride rich lipoproteins and related traits: A two-sample Mendelian randomization analysis

**DOI:** 10.64898/2026.04.27.26351885

**Authors:** Rehana Rasul, Mary Schooling, Ghada Soliman, Joy Shi, Zach Shahn

## Abstract

**INTRODUCTION:** Most randomized controlled trials (RCTs) found that omega-3 fatty acids have little to no effect on cardiovascular disease risk. However, a few suggested that a specific omega-3 fatty acid, eicosapentaenoic acid (EPA), reduces cardiovascular disease risk in patients with high triglycerides (TG). It is unclear whether EPA is beneficial in the general population or how it affects triglyceride-rich lipoproteins (TRL) and related traits. Using two-sample Mendelian randomization (MR), this study aimed to evaluate whether EPA has a protective effect on ischemic heart disease (IHD), TRL, and related traits in a general population.

**METHODS:** Associations of genetic variants with plasma EPA (EPIC-Norfolk, INTERVAL; n=14,267), and the outcomes IHD (Aragam et al., cases/n=181,522/1,165,690; FinnGen, N cases/n=31,640/218792), TRL, and related traits (Karjalainen et al.; n=68,559) were based on summaries from previous genome wide association studies (GWAS) of participants of European descent. Using eight proposed instruments associated with plasma EPA (P<5*10^−5^), inverse-variance weighted (IVW), MR-Egger, and weighted median (WM) estimators were used to determine the effect of a period shift in the natural log of plasma EPA one standard deviation, or EPA, on these outcomes.

**RESULTS:** Using IVW, EPA was associated with higher odds of IHD (OR=1.05; 95% CI=1.00, 1.10), but the CI included the null value. The WM estimate was similar, and the MR-Egger estimate was closer to the null (OR=1.01; 95% CI: 0.90, 1.11). EPA was associated with lower serum TG and lower large to small very low-density lipoprotein (VLDL) particle concentrations, but with increases in very small VLDL, intermediate density lipoproteins, and low-density lipoproteins. Although the distribution changed from larger to smaller TRL, there was no change in apolipoprotein B. EPA was also associated with increases in very large to medium high-density lipoprotein (HDL) particles and no change in small HDL, consistent with an increase in apolipoprotein A-I. EPA was also associated with increases in both remnant cholesterol and total serum cholesterol.

**DISCUSSION:** This study suggests that EPA may not have a beneficial effect on IHD in the general population of European ancestry. Rather, EPA appears to remodel TRL, possibly through lipolysis of large particles without full clearance of the resulting smaller particles, and this may have mixed implications for cardiovascular disease risk. A cardiovascular outcome trial of EPA monotherapy in a general population that collects lipid/lipoprotein subfractions would be needed to confirm these findings.

## INTRODUCTION

An estimated 239 million people were living with ischemic heart disease (IHD) in 2023, and this has increased due to changing risk factors, population growth, and population aging.^1,2^ Low-density lipoprotein (LDL) cholesterol causes atherosclerosis, and cholesterol-lowering therapies such as statins have been shown to reduce the risk of IHD among individuals with high risk of cardiovascular disease (CVD).^3,4^ Despite these medications, there remains high residual risk of CVD and adjuvant therapies are also needed.^5–7^ Because patients with high risk of CVD often present with high levels of triglycerides (TG), TG-lowering medications such as omega-3 fatty acids (OM3FA) were previously considered potential treatments. These fatty acids can be produced endogenously from consuming foods rich in alpha-linolic acid (Figure 1a), but are more efficiently obtained by directly consuming fish and fish oil supplements.^8^ Two OM3FAs, eicosapentaenoic acid (EPA) and docosahexaenoic acid (DHA), have been correlated with lower CVD incidence and mortality in cohort studies,^9,10^ but most randomized controlled trials (RCTs) found little to no improvement in CVD risk when using EPA and DHA in combination.^11–15^ This result was seen in trials using high risk CVD populations like the STRENGTH trial,^16^ as well as the VITAL trial, whose participants were healthy at baseline.^17^ These results were also consistent with null findings reported in subsequent Mendelian randomization studies of OM3FA.^18,19^

**Figure 1.**
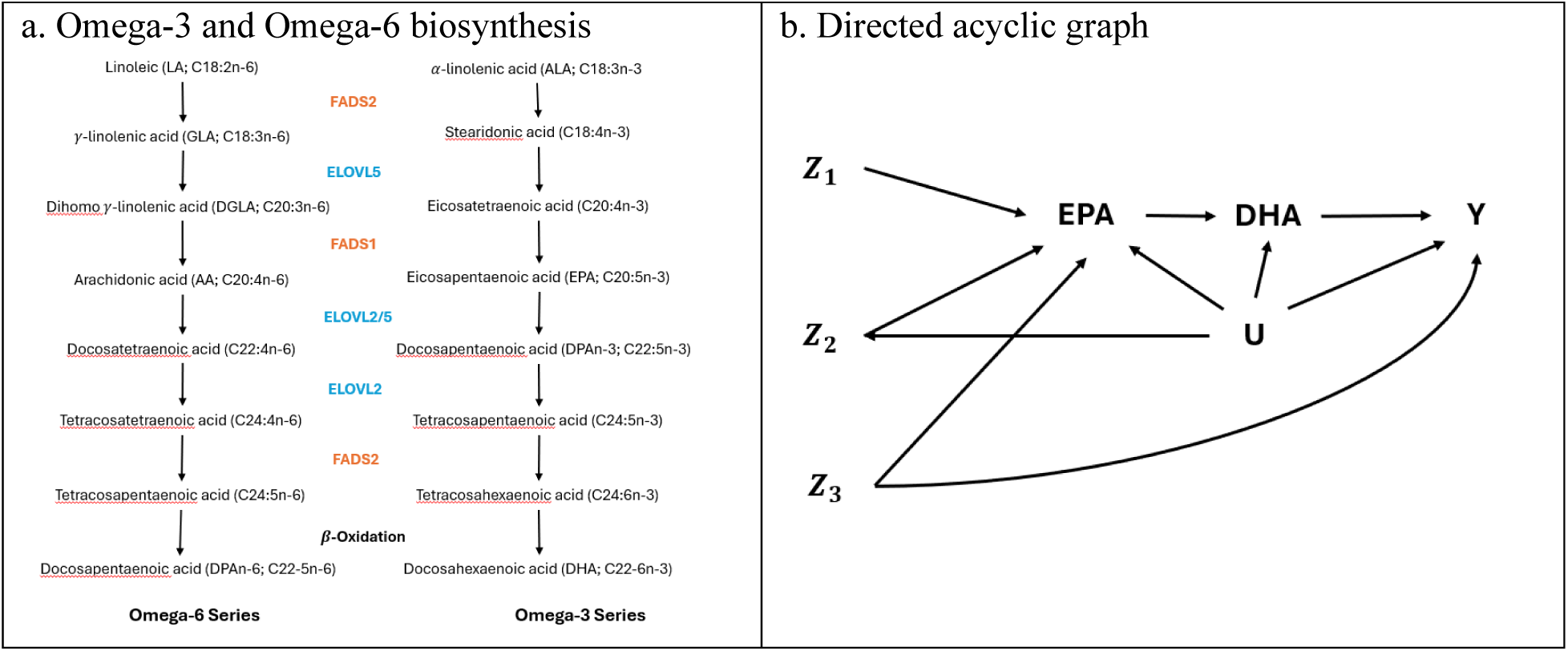
a. Omega-3 (OM3FA) and omega-6 (OM6FA) biosynthesis from fatty acid desaturase (FADS) and elongase (ELOVL) genes. b. Directed acyclical graph of hypothesized causal relationship between EPA and outcome Y. EPA is a sustained EPA treatment for plasma EPA across adulthood. Z_1_, Z_2_, and Z_3_ are vectors of genetic variants associated with EPA biosynthesis. U is a vector of unmeasured confounders. Y is an outcome (ischemic heart disease, triglyceride-rich lipoproteins and related traits, parental attained age). Variants in Z_2_ violate the exclusion restriction criteria and exchangeability assumptions of Mendelian randomization because they are associated with unmeasured confounders. Variants in Z_3_ violate the exclusion restriction criteria by causing Y through known pleiotropic paths like OM6FA synthesis or unknown paths. Variants in Z_1_ are valid instruments.

Several trials among patients at high risk for CVD found that EPA monotherapy reduced CVD risk compared to control for both primary and secondary prevention. In 2007, the JELIS trial found that EPA monotherapy was associated with 19% reduction in CVD incidence compared to low-intensity statins alone among Japanese participants^20^ and in 2019, the REDUCE-IT trial found that a purified form of EPA reduced hazards of CVD by 25% compared to mineral oil placebo.^21^ In 2024, the RESPECT-EPA trial also found 21% reduced risk of CVD in Japanese patients with established coronary artery disease.^22^ Meta-analyses suggested that EPA monotherapy may be more effective than mixed EPA/DHA on both CVD risk and CVD mortality.^14,15^ These studies signal that EPA and DHA may have different effects on CVD risk in high risk populations.

A possible mechanism by which EPA affects CVD is through triglyceride rich lipoprotein (TRL) particles, which transport TG and cholesterol esters (CE) through the blood.^23^ Chylomicrons, the largest TRL, are formed in the intestine and have the highest TG:CE content. Very low-density lipoproteins (VLDL) are the next largest and are synthesized in the liver. VLDL are lipolyzed by lipoprotein lipase (LPL) and hepatic lipase into smaller VLDL, intermediate density lipoproteins (IDL), and low-density lipoproteins (LDL). TRL are cleared out of circulation via the LDL receptor or LDL receptor related proteins. Both EPA and DHA alter the quantity and type of TRL by reducing TG, possibly through hepatic VLDL synthesis, lipolysis, or hepatic clearance.^24,25^ In the liver, EPA decreases VLDL biosynthesis and lipogenesis. EPA also increases TRL lipolysis through LPL expression.^26^ It is not clear how EPA affects TRL clearance directly. A reduction in VLDL synthesis, coupled with increases in lipolysis and clearance, is expected to shift the entire TRL distribution downwards and decrease atherogenesis.^25^ However, with inefficient lipolysis and inadequate clearance, the trajectory may shift from larger TRL to smaller TRL that may stay longer in circulation as remnant particles, making them more atherogenic.^25^ It is possible that EPA may promote TG reduction through efficient lipolysis, with compensatory TRL clearance, and that with DHA, lipolysis may lead to an increase in remnant particles and reduced clearance. A systematic review of short term randomized controlled trials (4-10 weeks) suggested that EPA had little effect on LDL, whereas DHA may increase LDL cholesterol and particle size using studies from both healthy and high risk populations.^27,28^ In cardiovascular outcome trials of high risk individuals, LDL decreased with EPA monotherapy compared to placebo,^21,22^ but in trials of mixed EPA/DHA compared to placebo, LDL was constant or increased and apoB was constant.^16^

EPA monotherapy in the high-risk CVD population appears promising, but it has not been tested in the general population. Prior RCTs also had limited data collection of TRL and related traits to understand lipid mechanisms. Pending an RCT to address these gaps, under certain assumptions, Mendelian randomization (MR) can be used to assess the causal role of EPA on these outcomes in the general population. MR is a form of instrumental variable analysis using genetic variants, which are randomly assigned conditional on parents’ genes, as proposed instruments to estimate the effect of an exposure affected by those variants on an outcome.^29,30^ This design is robust to unmeasured confounding between the exposure and the outcome, but relies on an alternative set of assumptions.^29,31^

Previous MR studies demonstrated no effect of plasma OM3FA or plasma DHA on IHD in general European populations.^18,19,32–35^ Few have evaluated EPA, and results have been mixed, possibly due to the instruments used.^19,34,35^ To be able to identify effects in MR, instruments cannot be horizontally pleiotropic, which occurs if they cause the outcome through pathways other than through the exposure.^30^ Methods robust to this assumption exist, but they make alternate assumptions and often require many instruments.^36^ Two studies used few genetic variants, which restricted the use of these methods.^34,35^ Using six instruments, Xu et al. found effects directionally different for most outcomes when using pleiotropy-robust methods compared to their main analysis which assumed that all instruments were not horizontally pleiotropic.^19^ Their result that EPA did not affect TG in their main analysis was also inconsistent with biological understanding,^37^ raising concerns that their MR assumptions may be violated. An updated MR analysis of EPA’s role on IHD, and expanding the outcomes to TRL and related traits, may provide some clarity, although a trial in the general population is preferable. Following the STROBE-MR guidelines,^38^ this study aims to use two-sample MR with summary data from genome-wide association studies (GWAS) to estimate the causal role of EPA on IHD, TRL, and related traits in the general population.

## METHODS

### Data sources

Separate data sources were used to select single nucleotide polymorphisms (SNPs), a type of genetic variant, to narrow the selection of candidate instrumental variables (selection datasets), provide estimates for the instrument-exposure associations (exposure dataset), and provide estimates for the instrument-outcome associations (outcome datasets). See Supplemental File 1 S1 for a description of data sources. Using separate datasets for selection and analysis is recommended to avoid bias due to winner’s curse.^39,40^ This bias can occur because associations based on p-value thresholds found in a discovery dataset may be overestimated due to the large number of tests performed. In the same dataset, associations between these variants and other correlated traits, including the outcome, may also be overestimated. Genetic variants were annotated using Ensemble and linked across datasets.^41^ Harmonization of the exposure and outcome datasets on the same effect allele was conducted using the *harmonise* function in the R package, *TwoSampleMR*.^42^

### Selection datasets: Candidate genetic instruments for plasma EPA

Summary associations of genetic variants with plasma EPA were obtained from a meta-analysis of GWAS studies from the Cohorts for Heart and Aging Research in Genomic Epidemiology (CHARGE) consortium (n=8,866; 100% European ancestry; age 18+) and from the Canadian Longitudinal Study of Aging (CLSA) (n=8,096; 100% European ancestry; age 45-85).^43–45^ For CHARGE, plasma EPA was measured as percentage of total fatty acids. Linear regressions were performed separately for each cohort and each adjusted for age, sex, and site of recruitment where appropriate. In addition, analyses from three cohorts within the CHARGE consortium (CARDIA, CHS and MESA) were adjusted for principal components. For CLSA, plasma EPA was natural log-transformed, trimmed to remove outliers, and standardized. Linear regressions were adjusted for age, sex, hour since last meal or drink, genotyping batch, and the first 10 genetic principal components.

### Exposure dataset: Associations of genetic instruments with plasma EPA

Summary statistics of the associations of SNPs with plasma EPA were obtained from a meta-analysis of GWAS studies from two UK-based prospective cohorts, EPIC-Norfolk (n=5,841; 100% European ancestry; age 40-79) and INTERVAL (n=8,455; 100% European ancestry; age 18+) conducted by Surendran et al (n=14,267) for the exposure dataset.^46–48^ For both cohorts, plasma EPA was natural log-transformed, winsorized, and standardized, adjusting for age and sex. For INTERVAL, linear regressions were adjusted for age, sex, center, the first five ancestry principal components, and sample processing variables (e.g., batch, plate number). For the EPIC-Norfolk cohort, linear regressions were adjusted for age, sex, and batch. Betas, interpreted as the average change in the standard deviation (sd) of the natural log of plasma EPA, adjusted for covariates, for the effect allele compared to the reference allele and standard errors (SE) were provided.

### Outcome datasets: Genetic associations with IHD

Summary data for associations of genetic variants with IHD were from a meta-analysis by Aragam et al. using data from the UK Biobank, the CARDIoGRAMplusC4D Consortium, and 9 other studies (cases/n=181,522/1,165,690; 100% European ancestry).^49^ The presence of IHD was defined differently for each study and may be prevalent or incident. Additive logistic or logistic mixed models with IHD status as the outcome were adjusted for study-specific covariates separately for each study, and all except the DECODE study also adjusted for ancestry principal components. Betas, interpreted as the change in the log-odds of IHD status for the effect allele compared to the reference allele, and SE were provided. It is noted that a small number of EPIC-Norfolk participants from the exposure dataset may also be in EPIC-CVD, one of the studies used for the genetic associations with IHD and this may lead to incorrect SEs. Genetic associations with IHD from the FinnGen cohort (European ancestry, N cases/n=31,640/218,792) were also used.^50^ The IHD definition included evidence of angina, MI, complications of MI, post-MI events, coronary atherosclerosis, and coronary revascularization from ICD-10 hospital discharge codes and cause of death registry. Summary statistics were from the IEU Open GWAS Project and extracted using the *TwoSampleMR* package.^42^

### Outcome datasets: Genetic associations with TRL and related traits

Summary associations of genetic variants with TRL measured in fasting participants were from a meta-analysis by Karjalainen et al. using data from 26 cohorts (N=68,559, 100% European ancestry).^51^ The NMR metabolomics platform was used to provide measures of lipoprotein subclasses, lipid concentrations, apoA1 and apoB, cholesterol, and triglyceride measures. Trait distributions were adjusted for age, sex, principal components, and study-specific covariates, and residuals were inverse rank normal transformed. Associations based on fasting participants were provided by the study authors. Betas, interpreted as the average change in the sd of the concentration or serum value of the trait, adjusted for covariates, for the effect allele compared to the reference allele, and SEs were provided.

### Outcome dataset for sensitivity analyses: Genetic associations with age at recruitment to the UK Biobank

To check for bias due to selection into the cohorts used in this study, genetic associations with age at recruitment in years to the UK Biobank (European ancestry, N= 361,194) from the Neale labs were evaluated.^52^ They were obtained from the IEU Open GWAS Project and extracted using the *TwoSampleMR* package (ukb-d-age).^42^ Although the age distribution for recruitment into studies from the data sources varied, their average ages were mostly within the age range for participants when they were recruited to the UK Biobank. See Supplemental File 1 Table S1 for their distributions.

### Data Analysis

#### Data generating mechanism

Figure 1b depicts a directed acyclic graph (DAG) of the hypothesized causal relationship between candidate instruments (Z_1_, Z_2_, and Z_3_), a sustained EPA level, and outcome Y. Z_1_, Z_2_, and Z_3_ are vectors of genetic variants associated with EPA biosynthesis., but only variants in Z_1_ satisfy the criteria to be valid instrumental variables, discussed below. The hypothetical EPA intervention of interest would shift the trajectory of the natural log of plasma EPA 1 standard deviation (sd) across the adulthood period for each individual. Such an intervention is not quite actionable with current technology but could be approximated via EPA supplements. Y is a binary or continuous outcome (IHD, TRL and related traits or age at recruitment, respectively). U is a vector of unmeasured confounders of the exposure-outcome relationship. For simplicity, the DAG includes only one time point, but it could be extended to have multiple treatment and outcome time points. It also does not show all possible mechanisms by which the exchangeability assumption could be violated (e.g., selection bias).

#### Causal estimand and identification assumptions

The causal effect of interest is a period effect that represents a contrast in the outcome under (1) no intervention, where EPA remains at its natural level and (2) a hypothetical intervention that increases EPA by one sd above its natural value throughout adulthood (see Supplemental File 2 S1).^53^ This effect will be expressed as a mean difference for continuous outcomes or an odds ratio for binary outcomes.^54^ For brevity, a one sd shift in the trajectory of the natural log of plasma EPA across the adulthood period is sometimes referred to simply as “EPA”. For MR, each genetic variant is a valid instrument if it meets three sufficient conditions: (1) it is associated with the exposure trajectory (relevance); (2) it does not affect the outcome except through the exposure trajectory (exclusion-restriction); and (3) it should not share common causes or other biasing paths with the outcome (exchangeability). In Figure 1b, Z_2_ includes genetic variants associated with confounders of the exposure-outcome relationships, such as smoking, alcohol use, and physical activity, violating the exchangeability assumption. Z_3_ includes genetic variants, such as those on the FADS and ELOVL gene clusters, which are associated with omega-6 fatty acids, creating a pathway between the variants and the outcome outside of the exposure, violating the exclusion-restriction. Only variants in Z_1_ satisfy the criteria to be valid instrumental variables.

The first condition, relevance, can be empirically verified—relevant variants can be identified from GWAS for one time point and are assumed to hold across the period. The other two conditions cannot be empirically verified. For example, while variants known to violate the exclusion restriction assumption can be removed, horizontal pleiotropy through unknown paths may still exist among the remaining variants. Similarly, exchangeability may be compromised by population stratification or by selection bias arising from misalignment in the timing of instrument assignment (at conception), assessment of eligibility for the MR analysis (which typically occurs decades later), and the start of follow-up. Here, we considered the use of alternative estimators, applied certain study restrictions, and used falsification strategies to address potential violations of these assumptions and assess their plausibility. We used the following robust MR estimators that relax or rely on alternative assumptions: (1) MR-Egger, which can be used to quantify directional horizontal pleiotropy and obtain unbiased effects in the presence of exclusion restriction violations if the Instrument Strength Independent of Direct Effect (InSIDE) assumption (i.e., pleiotropic effects are independently distributed from the instrument-exposure association);^55^ and (2) the weighted median (WM) estimator, which requires only half of the proposed instruments to be valid.^56^ We also restricted the analysis to a study population of the same ancestry, which mitigates concerns of bias from population stratification. We used TG and VLDL particle concentration, traits known to be reduced by EPA from RCTs^57^, as positive control outcomes to check instrument validity. Observed positive associations between EPA and these traits suggest that one of the assumptions may be violated. Lastly, we used age at recruitment as a negative control outcome—that is, because we would not expect EPA to affect age at recruitment, any observed association may be indicative of potential selection bias due to entry into the cohorts via survival or otherwise that would also arise when evaluating the effect of EPA on IHD. Of note, we expect survival bias to be minimal, as the mean age for most cohorts was middle aged (Supplemental File 1 S1), and therefore death from prior IHD or other causes is expected to be low.

To identify a period effect, additional assumptions are needed. Below we describe these assumptions for the continuous TRL outcomes. Modified versions of these assumptions appropriate for the odds ratio scale are needed for the estimators of the binary outcome. First, we assume homogeneity—that is, the effect of plasma EPA at a given time point is not modified by the instrument. This is often justified by asserting that there is no effect modification by confounder, conditional on the instrument and exposure trajectory.^58^ For example, diet can easily be a confounder and an effect modifier of this relationship. We also assume that there are no causal interactions between EPA levels at different time points. Furthermore, we assume the instrument-exposure association is constant on the additive scale over adulthood, since we only have one exposure measurement during this period. It is possible that conversion rates of ALA to EPA vary with age, especially for women, based on child-bearing and menopausal periods, which may invalidate this assumption.^59^ Linearity in the exposure-outcome association is also assumed. Linear models may possibly be mis-specified. When pooling effect estimates across multiple instruments, the effect should be homogeneous across variants. Cochran’s Q can be used to quantify heterogeneity of effects. Both samples should also arise from the same underlying population distribution, which is likely since the data were restricted to cohorts of European ancestry. Since several of these assumptions are difficult to justify contextually, although estimates were presented, the interpretation of presence of an average period effect rather than its magnitude is advised.

#### Identification of candidate genetic instruments

This study used a data-driven approach to select instrumental variables (Figure 2). First, variants associated with plasma EPA at P<5*10^−5^ from the selection datasets were included. When variants were in both datasets, p-values were based on associations from the CHARGE consortium. Second, since some variants in the selection dataset were not strongly associated with EPA in the exposure dataset, to avoid retaining weak instruments, variants were restricted to those also associated with plasma EPA at P<5*10^−5^ in the exposure dataset. Although this may introduce some bias due to winner’s curse, variants identified in both selection and exposure datasets would be unlikely due to chance. Third, variants which were palindromic, not biallelic, or had MAF≤0.01 were excluded. Fourth, known horizontally pleiotropic variants were excluded. These included variants on or correlated (R^2^>0.8) with variants on the *FADS* and *ELOVL* genes, which code enzymes to synthesize EPA, but also synthesize omega-6 fatty acids (linoleic acid, gamma-linolenic acid, dihomo-gamma-linoleic acid, arachidonic acid, stearidonic acid), which may affect the outcomes studied (i.e., variants in Z_3_ from Figure 1b). Variants are expected to be associated with DHA vertically since there is limited conversion of EPA to DHA, but variants with strong associations with DHA were also removed. Associations between variants and hypothesized confounders of the EPA-outcome relationships (smoking, alcohol use, education, physical activity) known to not be affected by EPA (i.e., variants in Z_2_ from Figure 1b) were also removed. These associations were identified from summaries in the NHGRI-EBI GWAS Catalogue obtained using the LDTrait tool.^60,61^ Fifth, the set was pruned to include only mildly correlated instruments (R^2^ ≤ 0.1, ≥ 10,000 kb) using PLINK called from the *TwoSampleMR* package.^42^ It used a greedy algorithm to identify variants in linkage disequilibrium and keep the one with the lowest p-value from the selection dataset. Sixth, TG and VLDL particle concentration were used as positive control outcomes to check instrument validity. Using estimates from the Wald estimator, variants demonstrating that EPA increased the positive control outcomes were excluded. An approximation to the F statistic in the exposure dataset was used to estimate the strength of each instrument.^62^

**Figure 2.**
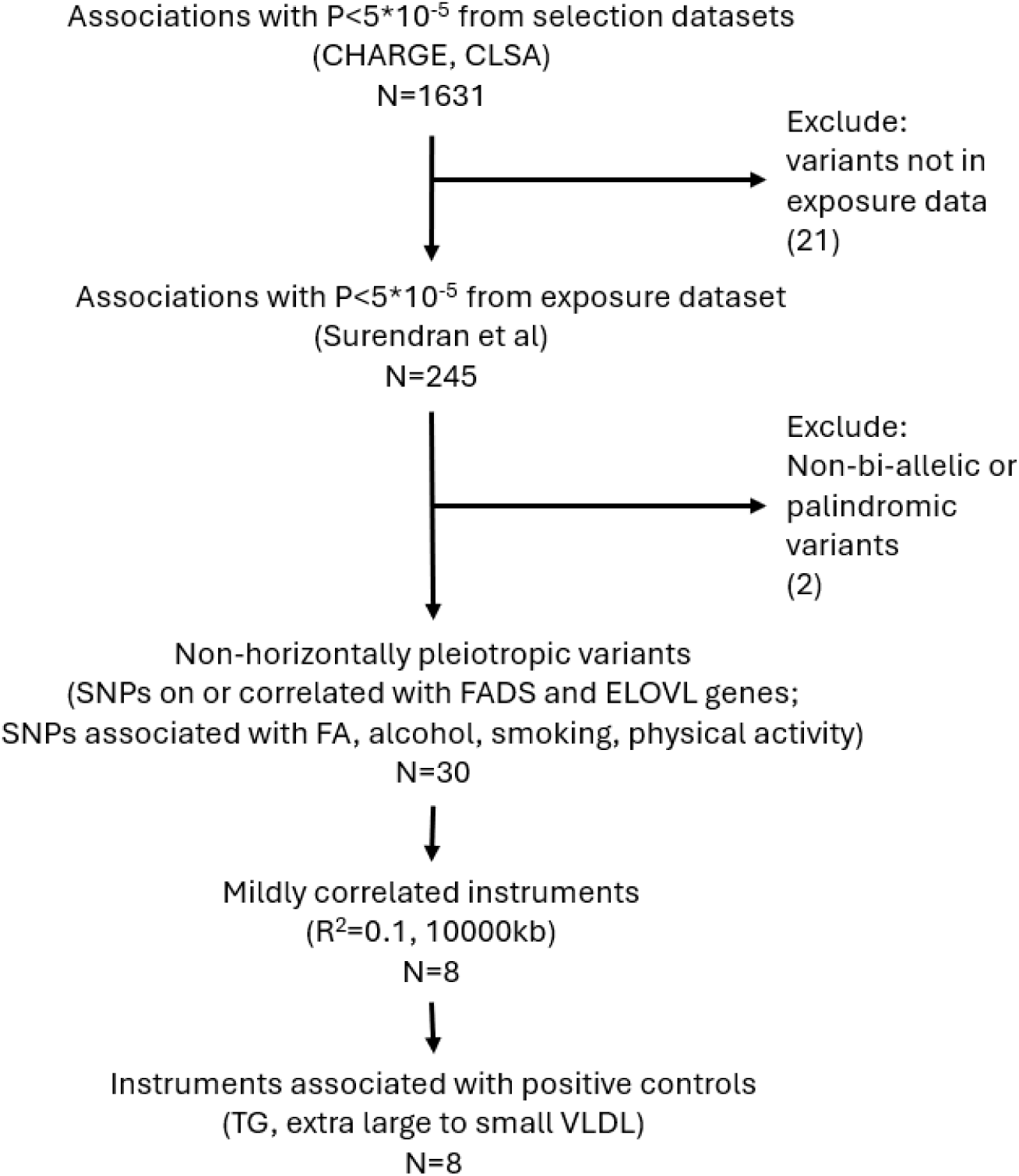
Flow chart to identify genetic variants for instruments. Selection datasets were from a meta-analysis of the cohorts from the Cohorts for Heart and Aging Research in Genomic Epidemiology (CHARGE) consortium and the Canadian Longitudinal Study of Aging (CLSA). The exposure dataset was from a meta-analysis of data using the EPIC-Norfolk and INTERVAL cohorts. FA=fatty acid; TG=triglycerides; VLDL=very low-density lipoprotein.

#### Estimators and estimation

For the main analysis, the inverse-variance weighted (IVW) estimator with multiplicative random effects, accounting for correlated instruments, was used to meta-analyze individual causal estimates based on the Wald ratio (see Supplemental File 1 S1).^63^ This method assumes linearity in the exposure-outcome association, homogeneity of this association by the instrument, and that all instruments are valid to yield unbiased estimates.^64^ Since there were two sets of genetic associations with IHD from different cohorts, separate MR analyses were performed for each. Additionally, genetic associations with IHD from both were pooled using fixed effects meta-analyses and MR was also conducted using these pooled associations. If a result had a Bonferroni-corrected significance level < 0.05/24 = 0.002 (24 outcomes), the effect of EPA on the outcome was considered to be statistically significant. Prior to analyses, alleles were oriented to represent increasing plasma EPA and this orientation was matched in the genetic correlation matrix. The *MendelianRandomization v. 0.10.0* package was used for analyses and the *ld_matrix* function from the *TwoSampleMR v. 6.16* package, was used to extract signed pairwise correlations between genetic instruments from the 1000 Genomes phase 3 reference panel for a population of European ancestry.^42,65,66^

#### Sensitivity analyses

Several sensitivity analyses were performed to assess assumptions of methods used. First, to test the robustness of the IVW results from violations due to directional horizontal pleiotropy, MR-Egger and WM estimators were also used.^55,56^ Second, since the FADS and ELOVL genes are biologically relevant, albeit pleiotropic, analyses were performed again including variants on these genes as instruments and directional pleiotropy was assessed. Third, analyses were repeated using the p<5*10^−8^ threshold for SNP-trait associations in the selection dataset only and also in both the selection and exposure datasets to evaluate using a more conservative threshold. Fourth, to check potential selection bias due to survival to cohort recruitment, the associations between plasma EPA and age at recruitment were also evaluated using MR.

## RESULTS

### Instruments

Of the 245 variants associated with plasma EPA in both the selection and exposure datasets, 8 were included as instruments (Table 1). Their F-statistics ranged from 26.16-177.64 (mean=56.29). Seven were on the genes (*SYT7* (n=1), *MYRF* (n=2), *RAB3IL1* (n=3), and TMEM258 (n=1) on chromosome 11. These genes are dominant in several regions across the body. *SYT7* and *RAB3IL1* are responsible for cell membrane repair including phospholipid binding, vesicle fusion and lipid transport.^67^ *MYRF* activates transcription of central nervous system myelin genes, but is also involved in hydrolase activity.^67^ These variants were associated with fatty acid measures generally, as well as phospholipids, diaglycerol, and mean corpuscular hemoglobin.^60,61^ *TMEM258* encodes a protein involved in glycolysation. The variants on *MYRF* and *TMEM258* were also associated with many different metabolites, including lipids and lipoproteins. One variant was on chromosome 6 upstream from the *ELOVL2-AS1* gene, which is involved in elongating the fatty acid chain. For each variant, an increase in EPA was associated with either a decrease or null effect in the positive control outcomes, TG and all sizes of VLDL except very small VLDL particles (Supplemental File 2 Figure S2). For each instrument, as VLDL particle size decreased, the estimate shifted further towards the null and right of the null in some cases.

**Table 1.** Characteristics of the SNPs associated with plasma EPA.

| SNP | Chromosome | Position | Candidate gene | Allele1 | Allele2 | EAF | Effect | SE | P-value |
| --- | --- | --- | --- | --- | --- | --- | --- | --- | --- |
| rs11230741 | 11 | 61351107 | SYT7/LOC105369331 | A | G | 0.2128 | 0.07735 | 0.01512 | 3.15E-07 |
| rs12665478 | 6 | 11080825 |  | A | G | 0.4184 | 0.06126 | 0.01197 | 3.05E-07 |
| rs174474 | 11 | 61672265 | RAB3IL1 | T | C | 0.7795 | 0.10770 | 0.01446 | 9.37E-14 |
| rs17764935 | 11 | 61664757 | RAB3IL1 | G | A | 0.0592 | 0.14940 | 0.02530 | 3.50E-09 |
| rs198460 | 11 | 61524974 | MYRF/MYRF-AS1 | A | G | 0.4827 | 0.07698 | 0.01183 | 7.69E-11 |
| rs2727266 | 11 | 61704334 | RAB3IL1 | A | G | 0.9312 | 0.13971 | 0.02321 | 1.76E-09 |
| rs412334 | 11 | 61560261 | TMEM258/FEN1/MIR611 | T | C | 0.165 | 0.11404 | 0.01591 | 7.59E-13 |
| rs422249 | 11 | 61639488 |  | C | T | 0.3441 | 0.16745 | 0.01256 | 1.59E-40 |

Note. SNP=single nucleotide polymorphism and is identified by rsid number; EAF=effect allele frequency. Effect is a beta coefficient, interpreted as the average change in the standard deviation (sd) of the natural log of plasma EPA, adjusted for covariates, for Allele1 compared to Allele 2. Allele 1 and 2 have been oriented such that effects represent an increase in plasma EPA measure. SE=standard error. Chromosome position number is based on GRCh37 build. Values are based on GWAS summary statistics from Surendran et al.<sup>48</sup>

### IHD

Using meta-analyzed IHD data, from the IVW estimates, a hypothetical sustained shift in the natural log of plasma EPA trajectory over adulthood by 1 sd was associated with higher odds of IHD (OR=1.05; 95% CI:1.00, 1.10) but the CI contained the null value (Figure 3). The MR-Egger intercept (B=0.01, P=0.355) did not indicate directional horizontal pleiotropy. WM estimates were similar. MR-Egger estimates were closer to the null and were less precise (OR=1.01; 95% CI: 0.90, 1.11). Cochran’s Q=7.30 (df=7; P= 0.398) did not indicate excess heterogeneity across variants. For individual variants (Supplemental File 2 Figure S3), all estimates were positive or close to the null value. It is also noted that all variants were also either positively associated with IHD or had estimates close to the null value. When removing individual variants from the analysis, estimates still indicated a weak positive association, but CIs included or were close to the null value (Supplemental File 2 Figure S4).

**Figure 3.**
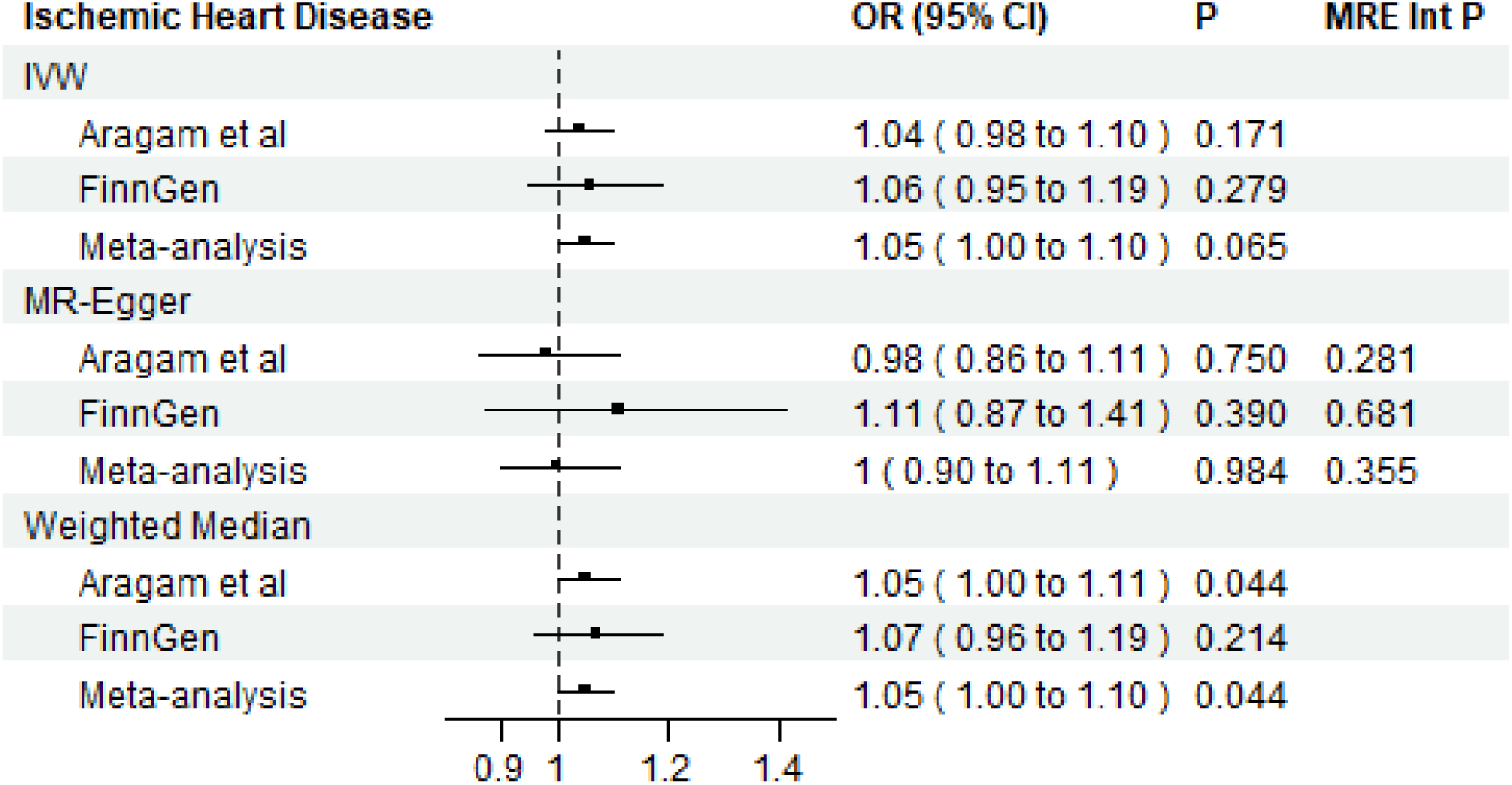
Univariable Mendelian randomization estimates for plasma EPA on ischemic heart disease. Results are expressed as change in odds of ischemic heart disease per standard deviation increase in the natural log of plasma EPA trajectory across adulthood and corresponding 95% confidence intervals (CI). OR=odds ratio; IVW = inverse-variance weighted; MR-Egger = MR-Egger; MRE Int P = P value for the intercept from the MR-Egger regression.

### Triglyceride rich lipoproteins

EPA was associated with lower mean diameter of VLDL particles (B= −0.26, 95% CI: −0.40, −0.12) (Figure 4). EPA was associated with bigger reductions of very large VLDL (B= −0.17, 95% CI: −0.27, −0.06) and this effect attenuated as VLDL particle size decreased to small VLDL (B= −0.11, 95% CI: −0.19, −0.03). A shift in the direction of effect was observed for very small VLDL (B= 0.11, 95% CI: 0.05, 0.17). EPA was also associated with increases in IDL (B=0.15, 95% CI=0.09, 0.21) and large (B=0.12, 95% CI: 0.07, 0.18) to small LDL (B=0.09, 95%CI: 0.04, 0.15). It was also associated with increased LDL particle diameter (B=0.09, 0.01, 0.17).

**Figure 4.**
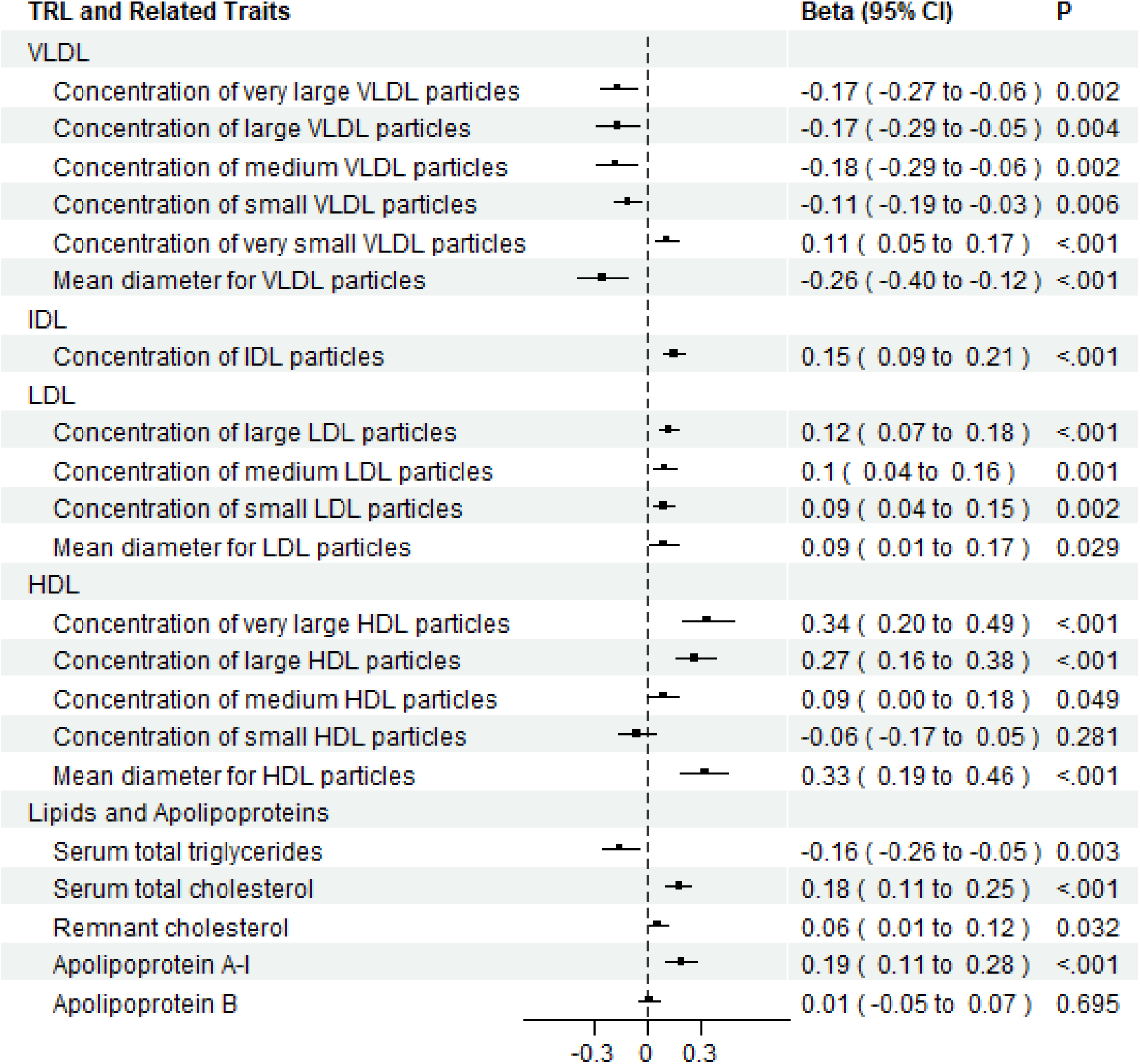
Univariable Mendelian randomization results for plasma EPA on triglyceride-rich lipoproteins and related traits among individuals of European ancestry. Results are expressed as beta coefficients of the change in triglyceride-rich lipoprotein per standard deviation increase in the natural log of plasma EPA trajectory across adulthood and corresponding 95% confidence intervals (CI) using the inverse-variance weighted method. TRL=triglyceride-rich lipoprotein; VLDL=very low density lipoprotein; IDL=intermediate density lipoprotein; LDL=low density lipoprotein; HDL=high density lipoprotein.

### HDL

EPA was associated with higher mean diameter for HDL particles (B=0.33, 95% CI: 0.19, 0.46). (Figure 4). The results suggest that EPA was not associated with small HDL (B=-0.06, 95% CI: −0.17, 0.05), but it was associated with higher amounts of larger HDL particles. It was associated with a 0.34 unit increase in very large HDL (B=0.34, 95% CI: 0.20, 0.49).

### Lipids and Apolipoproteins

EPA was associated with a reduction in TG (B=-0.16, 95%CI: −0.26, −0.05), but small increases in remnant cholesterol (B=0.06, 95% CI: 0.01, 0.12) and total serum cholesterol (B=0.18, 95%CI: 0.11, 0.25). It was not associated with a change in apoB (B=0.01, 95%CI: −0.05, 0.07), which is a measure of overall TRL particle number, but was associated with an increase in apoA-I (B=0.19, 95%CI: 0.11, 0.28), which is the primary structural apolipoprotein on HDL.

### Sensitivity Analyses

Estimates from the IVW method were similar to those from the WM method for all analyses. MR Egger estimates were closer to the null for IHD and stronger for TRL outcomes. MR-Egger estimates were also more imprecise, as expected since this method has lower power to detect effects.^55^ The MR-Egger intercept did not indicate unbalanced horizontal pleiotropy for the IHD analysis and for most lipid traits, except TG, VLDL, and HDL (Figure S3). When changing instrument selection criteria to include variants on the *FADS* locus and *ELOVL2* genes (n=7), EPA was associated with a small increase in odds of IHD and associated with a similar pattern of changes for TRL. When using the p<5*10-8 threshold for genetic variant-trait associations in the selection and exposure datasets (n=3) and only in the selection dataset (n=7), results were similar (Tables S3-S5). There was also no strong evidence of selection bias due to cohort recruitment based on the MR analysis with age at recruitment as a negative-control outcome (Figure S4).

## DISCUSSION

Using a two-sample MR design, this study provides evidence that EPA may not have protective effects on IHD in the general population during adulthood. An increase in EPA was associated with a reduction in TG and larger VLDL particles, as expected. However, EPA was also associated with increases in smaller TRL and larger HDL particles. Thus, increased EPA was associated with a shift in the distribution of TRL from larger to smaller particles, while having little effect on the total number of TRL particles.

Possible mechanisms to explain how EPA may affect IHD in the general population may be gleaned from the TRL pattern found in this study. Decreases in TG and larger VLDL particles due to EPA were expected from biological understanding.^57^ OM3FA are peroxisome proliferator-activated receptor (PPAR) agonists which reduce TG production by inhibiting VLDL biosynthesis in the liver. PPAR-alpha also activates LPL gene expression to promote metabolism of larger VLDL particles to smaller TRL.^68^ However, there was no indication of increased clearance, as evidenced by the lack of an association with apoB. The implications of stimulating LPL without also promoting compensatory LDL-R clearance on CVD risk are not clear. If smaller TRL are not transitory and stay in circulation as remnant particles, they may become more enriched with CE and, consequently, more atherogenic. However, the TG content from large VLDL has been associated with small, dense LDL, which are highly atherogenic.^25^ Promoting large VLDL metabolism may lead to larger, more buoyant LDL particles, which are thought to have a higher affinity for the LDL-R than smaller ones.^25^ There was an increase in LDL diameter, but it is noted that both large and small LDL particles increased. Implications of CVD risk due to EPA’s effect on HDL is also unclear. The increase in HDL particle size may indicate reverse cholesterol transport or cholesterol ester transfer protein exchange, which may also have different effects on CVD risk. This also affects both total and remnant cholesterol levels. Both increased due to EPA in this analysis, but the effect on remnant cholesterol was small.

The finding that EPA does not reduce CVD risk in a general population may be due to these potentially counteracting mechanisms through TRL. These results are also consistent with other MR studies,^19,34^ but they are different from EPA monotherapy trials showing that EPA monotherapy reduced CVD risk among participants with high CVD risk.^20–22^ Previous short term RCTs studying biological mechanisms and EPA monotherapy trials also found either no change or decreases in LDL, consistent with their reports of lower apoB number, but inconsistent with this study’s results.^20,21,28,69^ One possible explanation for the lack of benefit in CVD risk in this study compared to the trials is that the effect may be heterogeneous by CVD risk status or region. EPA may be protective for IHD in high-risk CVD populations, but not in the general population from the cohorts used in this analysis. Among high risk participants, stratified analyses from the REDUCE-IT trial found that EPA was more beneficial for secondary prevention participants than primary prevention patients.^21^ Effects may also be heterogeneous by region. All of the EPA monotherapy trials except the REDUCE-IT trial were conducted on Japanese participants.^20–22^ A post-hoc analysis of a mixed EPA/DHA trial of high-risk CVD participants also found favorable effects among Asians only.^16,70^ A second possible explanation is that effect heterogeneity by CVD risk status is not present and EPA monotherapy may not have a protective effect on IHD even in the trial populations. Although some have questioned the use of a mineral oil placebo in the REDUCE-IT trial,^71–73^ corroboration from the other two trials makes this explanation less likely. A third possible explanation is that the results of this MR study are biased and that EPA may be protective in both the general population and a high-risk CVD population. The positive control finding in this study that EPA reduces TG (in contrast to the results of Xu et al., ^19^ which may have been pleiotropic due to inclusion of variants from the FADS locus region) might be construed as evidence against a general bias toward harmful effects driving these results. Furthermore, the consistency of the array of lipid results with a simple mechanism of lipolysis without clearance might be perceived to support credibility (at least in the general population) via an Occam’s razor type argument. However, MR studies rely on strong, mostly unverifiable assumptions. None of these explanations can be logically ruled out and a trial of EPA monotherapy in the general population can clarify these effects.

Although some have suggested that EPA acts differently on CVD risk than DHA,^74^ the effect of EPA on CVD risk in this study was similar to effects from other MR studies evaluating DHA and combined OM3FA on CVD risk in general populations of European descent,^18,19,34^ and from a trial of mixed EPA/DHA treatment in a general US population.^17^ These findings suggest that EPA and DHA have similar effects on CVD risk in a general population, but there may be heterogeneous effects due to CVD risk status or region. In high-risk CVD populations, EPA monotherapy appears to be beneficial compared to mixed EPA/DHA, whereas trials evaluating mixed EPA/DHA had null results.^14,15^ However, since mixed EPA/DHA was also found beneficial among Asians with high-risk CVD^70^ and most of EPA monotherapy trials were conducted on Japanese participants, it is possible that differences in the mixed EPA/DHA trials and EPA monotherapy trials may not be due to their differences in the supplements evaluated, but due to region. Further research to investigates these potential effect modifiers is necessary to understand these effects.

Several strengths of this analysis should be noted. First, in lieu of an RCT, with an MR study design, given assumptions discussed above, results can be interpreted as causal even in the presence of unmeasured confounding. Second, this is the first MR analysis using data from Surendran et al. to study EPA,^48^ which allowed for comparison with previous results based on exposure data from the CHARGE consortium, as well as the ability to use separate selection and exposure studies for instrument identification. Effects of EPA on IHD were also not previously estimated using the FinnGen dataset. Third, this study investigated an expanded set of lipid traits collected during a fasting state. Combining the relative effect estimates on a range of lipids with biological theory enabled formulation of hypotheses regarding possible mechanisms of EPA on metabolic diseases. Fourth, using positive controls as part of instrument identification and interpretation of results strengthen these conclusions. Finally, the MR-Egger and WM results provide some reassurance that the exclusion-restriction was not seriously violated, although this is still an unverifiable assumption.

This study should be also interpreted with caution for several reasons. First, the core IV assumptions alone only license interpretation of results as tests of the null causal effect and not as estimates of its magnitude. The additional linearity and homogeneity assumptions required for estimation of effect magnitudes are very unlikely to hold. Second, two of the core assumptions, the exclusion restriction and the exchangeability assumptions, are unverifiable and here may be false. The MR-Egger estimator is only robust to the exclusion restriction if the InSIDE assumption is met and it also has lower power to detect effects, as shown by the larger confidence intervals. WM assumes at least half of standardized inverse-variance weights are from valid instruments. A violation of exchangeability due to selection bias is possible, although biases from survival into the cohort and competing diseases with shared etiology with the outcome are minimal for IHD. This study also did not evaluate sex-specific effects and selection bias may be stronger on effects among males, who have an earlier onset of IHD compared to females.^75^ Third, a data-driven approach for instrument identification was used, although use of instruments with known biological roles on the exposure are optimal.^76^ However, we did exclude candidates based on biological considerations. Fourth, use of pooled summary data from multiple studies with differences in study design and analysis may introduce bias in these estimates.^77^ Fifth, other important cardiovascular markers were not assessed. Chylomicrons and their remnants are difficult to study given their variability in timing of eating. Other characteristics of TRL, like plasma residence time and cholesterol cargo are also important in understanding atherogenic effects. Other apolipoproteins (e.g., apoC-II, apoC-III, apoE), angiopoietin-like proteins, and insulin, which are involved in TRL metabolism, may also clarify these pathways. Lastly, the MR analysis uses a general population of European ancestry which includes both healthy and unhealthy participants and may not be generalizable to other populations or certain sub-populations, such as people with heart disease.

## CONCLUSION

This Mendelian randomization study provided evidence that EPA may not be not protective against IHD in a general European ancestry population. It found that EPA has a strong effect on TRL remodeling, decreasing levels of large TRL particles and increasing levels of smaller particles. This remodeling is consistent with an explanation of reduced VLDL biosynthesis and increased lipolysis without adequate clearance. These effects have mixed implications for atherogenesis. A cardiovascular outcome trial with EPA and DHA monotherapy arms in a general population which also collects lipoprotein subfractions would be needed to understand the efficacy and mechanisms of OM3FA.

## ETHICS APPROVAL

This study was not considered human subjects research by the IRB of our institution. This study uses genetic summary statistics previously collected and made available publicly or can be requested from the data sources listed in the Data Availability Section. Materials involved in generating summary statistics from each data source were collected by the original cohorts with informed consent from participants.

## DATA AVAILABILITY

Summary statistics necessary to replicate the MR analyses are included in Supplemental File 1. Full GWAS data for plasma EPA from the CHARGE consortium (accession no: GCST001178) and the Canadian Longitudinal Study of Aging (accession no: GCST90200349), as well as for IHD from Aragam et al. (accession no: GCST90132314) are publicly available through the NHGRI-EBI GWAS catalogue^78^ (https://www.ebi.ac.uk/gwas/studies). Summary statistics for plasma EPA from Surendran et al. are publicly available through the interactive database https://omicscience.org/apps/mgwas^48^. Summary statistics for IHD from FinnGen and age at recruitment from the UK Biobank are publicly available through the IEU Open GWAS Project^42,79^ (https://gwas.mrcieu.ac.uk/) and can be extracted using the *TwoSampleMR* package (finn-b-I9_IHD; ukb-d-age). Full GWAS data for TRL under fasting conditions can be requested from Karjalainen et al.^51^

## ACKNOWLEDGEMENTS

We would like to acknowledge Surendran et al. for providing summary GWAS data for the plasma EPA trait and Karjalainen et al. for providing summary GWAS data for fasting-specific TRL and related traits.^48,51^ We also would like to acknowledge the time and effort of all study participants and investigators whose data has enabled this research for each of the cohorts.

