## Supplemental File 2 for "Causal role of EPA on ischemic heart disease, triglyceride rich lipoproteins and related traits: A two-sample Mendelian randomization analysis"

### SUPPLEMENTARY FILE 2

Rehana Rasul[1], Mary Schooling[1], Ghada Soliman[1], Joy Shi[2], Zach Shahn[1,3]

1. CUNY Graduate School of Public Health and Health Policy, New York, NY, United States
2. Department of Epidemiology, Harvard T.H. Chan School of Public Health, Boston, MA, United States
3. Institute for Implementation Science in Population Health, New York, NY, United States

**Corresponding Author**
Rehana Rasul
Department of Epidemiology and Biostatistics, Graduate School of Public Health and Health Policy, City University of New York, 55 West 125th Street, New York, NY 10027, US


**S1. Analysis considerations**

**1. Explanation of the causal estimand**

Let $a_t$ be EPA level at a particular time *t* (in years), where *t* is in the period of adulthood ranging from year *m-p* through $m$. $Y^{a_{m-p},\dots,a_m}$ is the counterfactual outcome had an individual received EPA levels $a_{m-p}$ through $a_m$ over this period and $E[Y^{a_{m-p},\dots,a_m}]$ is the mean counterfactual outcome had everyone in the population received these EPA levels over this period.[1] For continuous outcomes, the causal contrast of interest is $E[Y^{a_{m-p}+1,\dots,a_m+1}] - E[Y^{a_{m-p},\dots,a_m}]$, assumed constant over $(a_{m-p}, \dots, a_m)$, or the average period effect of shifting the trajectory of the natural log of plasma EPA by one sd across this period. For the binary outcome, the causal estimand of interest is an odds ratio.[2] For brevity, a one sd shift in the trajectory of the natural log of plasma EPA across the adulthood period is sometimes referred to simply as "EPA".

**2. Approximation to the F statistic**

An approximation to the F statistic,[3] $F_j \approx \frac{\hat{B}_{X_j}{}^2}{(se(\hat{B}_{X_j}))^2}$ in the exposure dataset was used to estimate the strength of each instrument, where $\hat{B}_{X_j}$ is the beta coefficient of the change in the sd of the natural log of plasma EPA due to SNP $j$.

**3. Description of the Estimators and Estimation Method**

For the main analysis, the inverse-variance weighted (IVW) estimator with multiplicative random effects, accounting for correlated instruments, was used to meta-analyze individual causal estimates based on the Wald ratio, $IVW_{correlatedIV} = (\hat{\beta}_X^T\,\Omega^{-1}\hat{\beta}_X)^{-1}(\hat{\beta}_X^T\,\Omega^{-1}\hat{\beta}_Y)$.[4] This method assumes linearity in the exposure-outcome association, homogeneity of this association by the instrument, and that all instruments are valid to yield unbiased estimates.[5] $\hat{\beta}_X$ is the vector of coefficients representing a one sd increase in the natural log of plasma EPA per reference allele for each instrument. $\hat{\beta}_Y$ is the vector of coefficients representing the change in log odds of IHD, or unit of TRL measure per reference allele for each instrument for IHD and TRL outcomes, respectively. For IHD, it is noted that the IV estimand based on the Wald ratio is an approximation to the population odds ratio.[2] It is also noted that covariate sets used for adjustment in the exposure and outcome datasets were not identical, but in most cases, each adjusted for age, sex, and ancestry principal components. $\Omega_{j_1 j_2}$ is a weighting matrix based on the standard errors of $\hat{\beta}_X$ and $\hat{\beta}_Y$ and the correlation $\rho_{j_1 j_2}$ between the instruments, with $\Omega_{j_1 j_2} = se(\hat{B}_{Yj_1})se(\hat{B}_{Yj_2})\rho_{j_1 j_2}$ for instruments $j_1$ and $j_2$.[4,6] Since there were two sets of genetic associations with IHD from different cohorts, separate MR analyses were performed for each. Additionally, genetic associations with IHD from both were pooled using fixed effects meta-analyses and MR was also conducted using these pooled associations. If a result had a Bonferroni-corrected significance level < 0.05/24 = 0.002 (24 outcomes), the effect of EPA on the outcome was considered to be statistically significant. Prior to analyses, alleles were oriented to represent increasing EPA and this orientation was matched in the genetic correlation matrix. The *MendelianRandomization v. 0.10.0* package was used for analyses and the *ld_matrix* function from the *TwoSampleMR v. 6.16* package, was used to extract signed pairwise correlations between genetic instruments from the 1000 Genomes phase 3 reference panel for a population of European ancestry.[7-9]

**Figure S2. Positive control outcomes.** Univariable Mendelian randomization results for plasma EPA on the positive control outcomes (serum triglycerides and very large to very small very low-density lipoproteins) per genetic variants among individuals of European ancestry. Results are expressed as beta coefficients of the change in each outcome per standard deviation increase in the natural log of plasma EPA trajectory across adulthood and corresponding 95% confidence intervals (CI) using the inverse-variance weighted method. TG=triglycerides; VLDL=very low-density lipoprotein; IVW=inverse-variance weighted.s

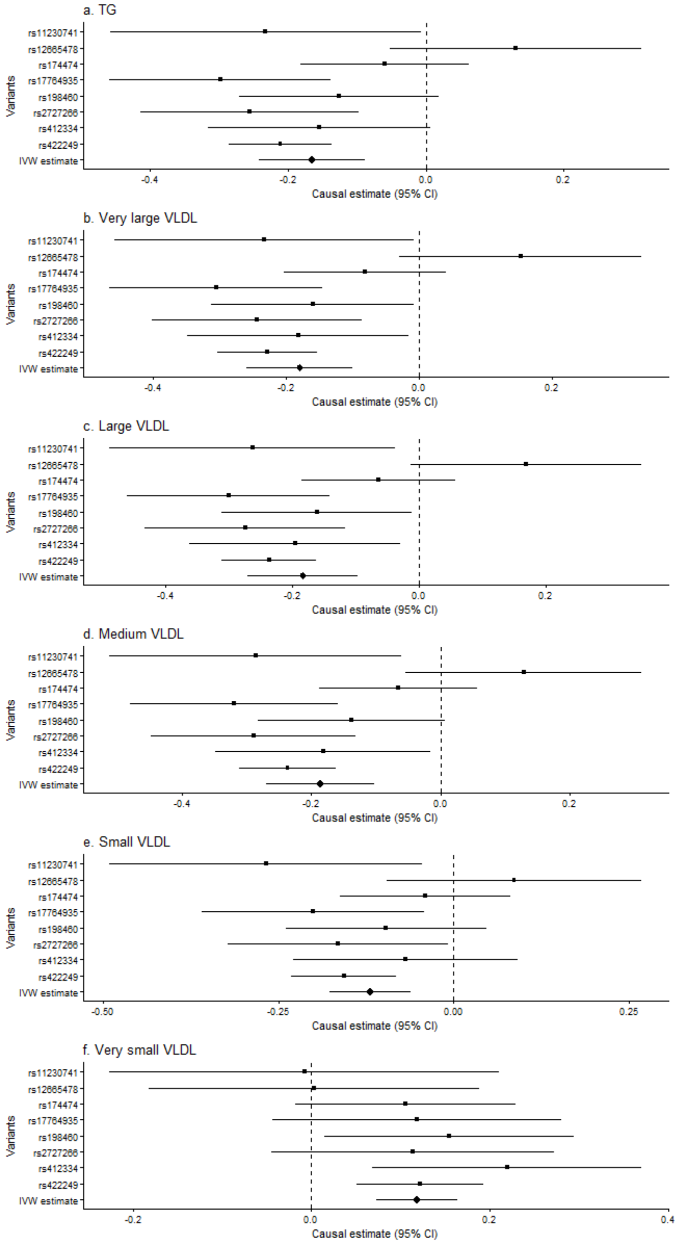

**Figure S3. Instrument-specific results for IHD.** Univariable Mendelian randomization estimates for plasma EPA on ischemic heart disease using data from (a) meta-analyzed data from Cardiogram and FinnGen, (b) Cardiogram, and (c) Finngen. Results are expressed as change in log odds of IHD per standard deviation increase in plasma EPA trajectory across adulthood and corresponding 95% confidence intervals (CI). IHD=ischemic heart disease; IVW = inverse-variance weighted.

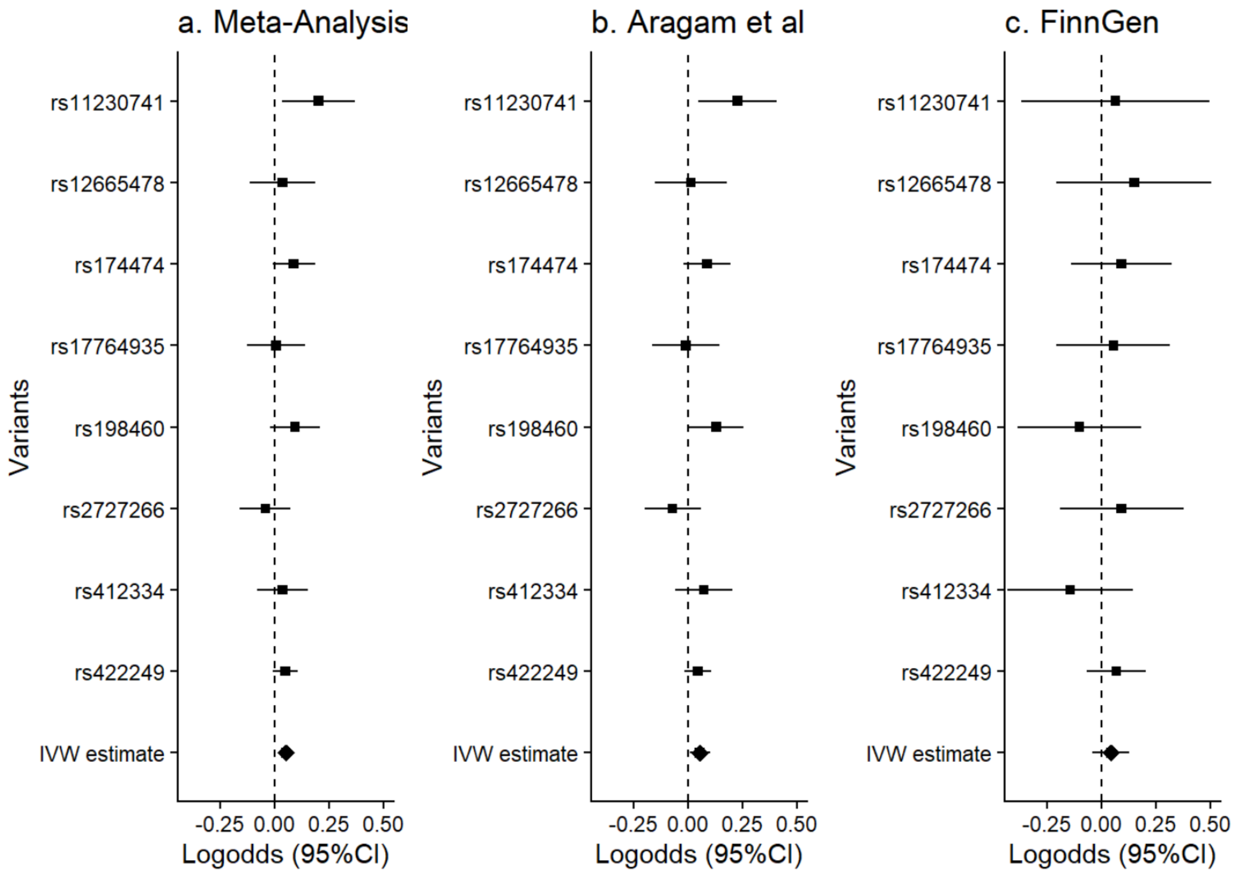

**Figure S4. Leave one out analysis for IHD.** Univariable Mendelian randomization estimates for plasma EPA on ischemic heart disease using data from (a) meta-analyzed data from Cardiogram and FinnGen, (b) Cardiogram, and (c) Finngen using all instruments except the one specified. Results are expressed as change in log odds of IHD per standard deviation increase in plasma EPA trajectory across adulthood and corresponding 95% confidence intervals (CI). IHD=ischemic heart disease; IVW = inverse-variance weighted.

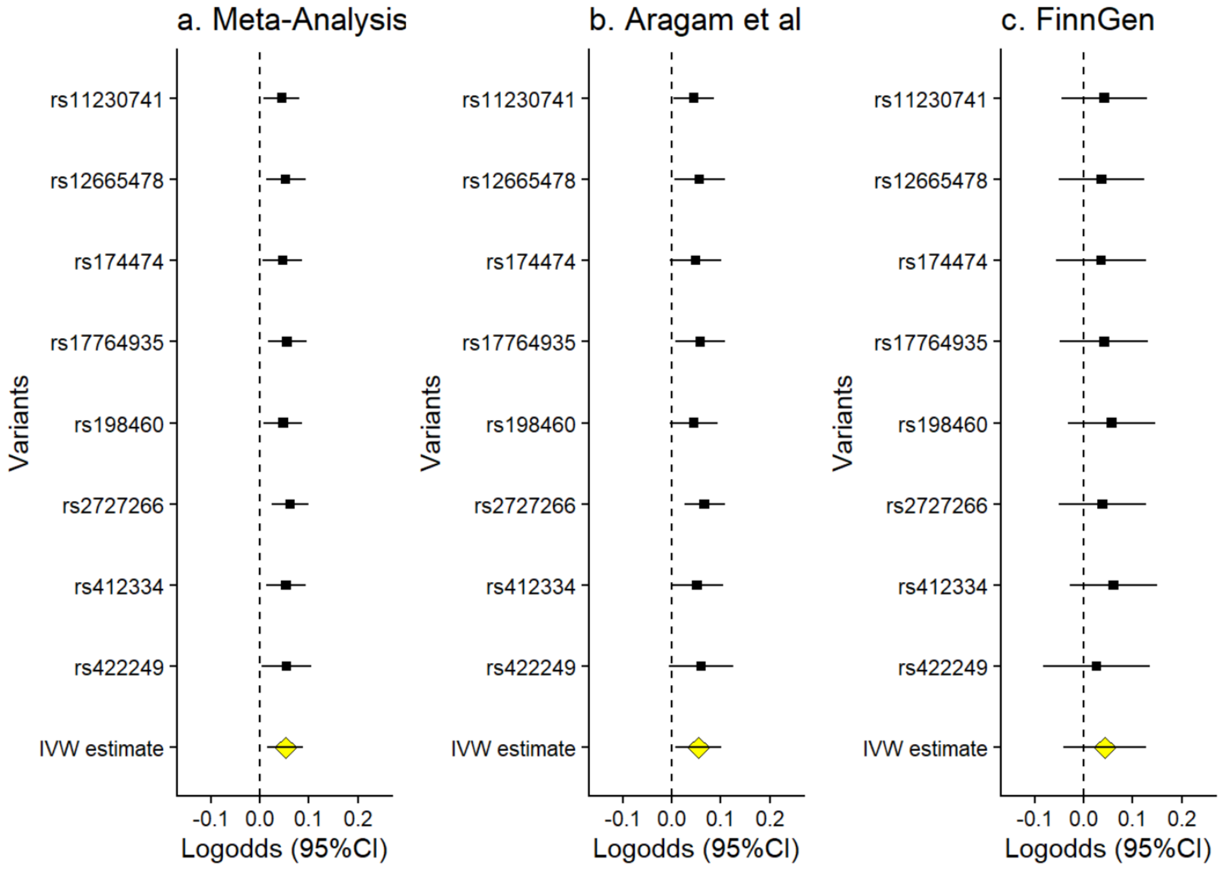

**Figure S5.** Univariable Mendelian randomization results for plasma EPA on age at recruitment to the UK Biobank. Results are expressed as beta coefficients of the change in age in years per standard deviation increase in the natural log of plasma EPA trajectory across adulthood and corresponding 95% confidence intervals (CI). IVW = inverse-variance weighted; MR-Egger = MR-Egger; WM=weighted median. MRE Int P = P value for the intercept from the MR-Egger regression.

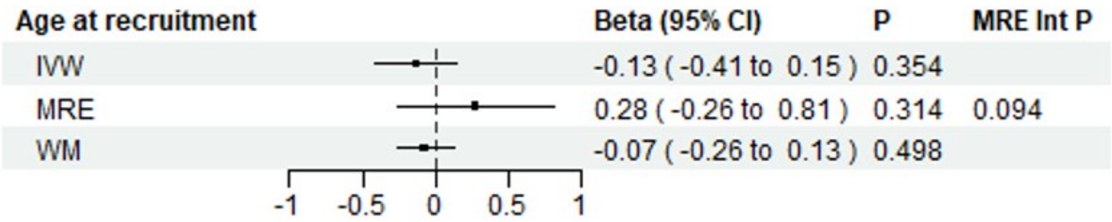
